# Health System Resilience-enhancing Strategies for Managing Public Health Emergencies of International Concerns (PHEIC): Success and Challenges A Systematic Review Protocol

**DOI:** 10.1101/2022.06.14.22276386

**Authors:** R. M. Nayani Umesha Rajapaksha, Resham Khatri, Chrishantha Abeysena, Millawage Supun Dilara Wijesinghe, Toms K. Thomas, Aklilu Endalamaw, Nadeeka Perera, Mekala Fernando, Gayani De Silva, Roshan Rambukwella, Yibeltal Assefa

## Abstract

**Introduction:** Health system resilience is the ability to prepare, manage, and learn from a sudden and unpredictable extreme change which impacts health systems. Health systems globally have recently been affected by a number of catastrophic events, including natural disasters, and infectious disease epidemics. Understanding health system resilience has never been more essential until emerging global pandemics. Therefore, the application of resilience-enhancing strategies with existing frameworks needs to be assessed to identify the management gaps and give valuable recommendations from the lessons learnt from the global pandemic.

**Methods:** The systematic review will be reported using the Preferred reporting items for systematic review and meta-analysis protocols guideline (PRISMA-P). Reporting data on health system building blocks and systematic searches on resilience enhancing strategies for the management of Public Health Emergencies of International Concerns (PHEIC) after the establishment of International Health Regulations (IHR) since managing PHEIC after the establishment of IHR in 2007 will be included. The search will be conducted in PubMed, Scopus, Web of Science, and Google Scholar.

**Discussion:** Health system resilience is key to coping with catastrophic events, such as the economic crisis and COVID-19 pandemic. The mapping of available literature towards the application of resilience-enhancing strategies with existing frameworks needs to be assessed to identify the management gaps and give valuable recommendations from the lessons learnt from the global pandemic to improve the level of preparedness and response to similar public health emergencies in the future.

**Conclusion:** A protocol for a global review of health system resilience for pandemic management is described. This review will add to the body of knowledge about health systems enhancing research and policy formulation.

**Strengths and limitations of this study Strengths:**

- Assessing success and challenges of resilience-enhancing strategies to identify the management gaps and give valuable recommendations from the lessons learnt from the global pandemic in future disasters
- The systematic review will be reported using the Preferred reporting items for systematic review and meta-analysis protocols guideline (PRISMA-P).
- Collaboration between Asia-Pacific researchers with different background

**Limitation:**

- Unable to do meta-analysis due to nature of the selected scope of the research

## Introduction

Health system resilience is defined as ‘the ability to prepare, manage and learn from a sudden and unpredictable extreme change which impacts on health systems’,[1]. It focuses on the health system preparedness and response to an emergency and coping capacity for changes following a crisis in view of absorbing, adapting, and transforming,[1-2]. However, the focus of the health system resilience was broadened with the improving number of literatures around health systems resilience, which extends to focus on vulnerability, system strains and everyday resilience,[2]. Understanding health system resilience has never been more essential until emerging global pandemics. Health systems globally have recently been affected by a number of catastrophic events, including natural disasters, infectious disease epidemics such as Ebola outbreak, novel coronavirus disease-19 (COVID-19) pandemic. Due to the new emergence and rapid transmission of the COVID-19 since late 2019, global health systems are severely affected. As stated by the WHO, all the organizations, institutions, resources, and people whose primary purpose is to improve health is included in the health system. Moreover, efforts to influence determinants of health and direct health-improvement activities are considered as the component of the health system. A health system delivers preventive, promotive, curative, and rehabilitative interventions combined with public health actions and the pyramid of healthcare facilities that provide personal healthcare by multi-stakeholders,[3]. A health system needs human resources, health finances, information, supplies, transport, communications, overall guidance, and direction to provide a quality service. Each area of the health system needs to be strengthened to address the key constraints. Therefore, the WHO described six building blocks of the health system framework which include ‘health service delivery, health workforce, health information systems, access to essential medicines/vaccines, health financing, leadership, and governance. These building blocks are targeted to achieve impacts, including improved health outcomes, equity, social and financial risk protection, responsiveness, and efficiency, which contribute to strengthening health systems to provide sustainable quality, efficient and effective health services,[3]. Furthermore, the need for understanding of governing of health systems is highlighted following catastrophic events occurring over the last decade. The majority of low- and middle-income countries were highly affected by the emergence of infectious diseases (e.g., Ebola, COVID-19) and armed conflicts,[4-6]. Moreover, managing the crises, national responses across the globe were varied, some focus on the transmission of diseases and prevention of deaths. Importantly, the countries having established good health systems also struggled to cope with the exponentially accelerating number of cases,[7]. With the exponential increase of pandemics, the International Health Regulations (IHR) was established in 2007 for governing the global health security. The IHR declared Public Health Emergencies of International Concern (PHEIC) Six event of PHEIC were documented between 2007 and 2020, including ‘H1N1 influenza pandemic (2009), Ebola (West African outbreak 2013–2015, outbreak in Democratic Republic of Congo 2018–2020), poliomyelitis (2014 to present), Zika (2016) and COVID-19 (2020 to present)’. Among them Poliomyelitis is the longest PHEIC, and Zika was the first PHEIC for arboviral disease. Even though the public health impact of the event was considered serious and associated with a potential for international spread, several other emerging diseases were not declared as PHEIC,[6]. Moreover, response measures following a crisis, improving surge capacity, and planning for further action to minimise vulnerability can be identified when assessing the health systems’ resilience,[8]. The purpose of assessing resilience and its interpretation depends on multiple contextual factors that need to be evaluated for better understanding. The well-assessed areas can be utilized as a starting point and encourage policymakers to adopt an appropriate strategy in a particular country. Furthermore, policymakers need to regularly review their health systems to assess their resilience,[9]. In addition, the application of resilience-enhancing strategies is new. The concept has become relevant and more researched with societal response to health emergencies during the past two decades,[10].

### Justification (Rationale)

Health system resilience is key to coping with catastrophic events, such as the economic crisis and COVID-19 pandemic. However, there is confusion about the meaning of resilience, strengthening it, and assessing it,[1]. Furthermore, health systems resilience research has reached a crucial point; health system interventions have only partly been followed by empirical research and concrete applications,[11]. Therefore, the application of resilience-enhancing strategies with existing frameworks needs to be assessed to identify the management gaps and give valuable recommendations from the lessons learnt from the global pandemic. Importantly, improving resilience could help the health system respond to a pandemic like COVID-19. However, most of the research has so far remained primarily theoretical. Therefore, applied research towards a cohesive set of goals needs to be identified and promoted to develop and implement strategies to strengthen systems. Furthermore, strengthening health systems resilience can be addressed during the pandemic and future disasters if the issues are correctly addressed during research and interpretation of findings,[11]. Therefore, there is a need to analyze the implementation of resilience-enhancing strategies to draw lessons from health systems that have proved more successful at dealing with the past crisis and offer evidence on best practices for health systems under strain. Moreover, it will invariably help improve the level of preparedness and response to similar public health emergencies in the future.

### Objectives

The objectives of this study are;

1. To identify the health system resilience-enhancing strategies for managing Public Health Emergencies of International Concerns (PHEIC) globally.
2. To describe the implementation of health system resilience-enhancing strategies for managing pandemics.
3. To identify success, opportunities, and challenges, and provide recommendations towards successful management of PHEIC.

### Review question

What are the success strategies, and challenges toward health system resilience for managing Public Health Emergencies of International Concerns (PHEIC) globally?

## Methods

### Type and method of review

Health system resilience: A Systematic review

### Study Period

30 ^th^ of March 2022 to 15 ^th^ October 2022

#### Anticipated or actual start date for the review

15 ^th^ of July 2022

### Anticipated or completion date

15 ^th^ of October 2022

### Eligibility criteria

#### Inclusion criteria

We intend to include all studies in English with any peer reviewed articles under the World Health Organization’s (WHO) six building blocks of health system that adhered to resilience enhancing strategies for the management of PHEIC after establishment of IHR since managing PHEIC after the establishment of IHR in 2007. Descriptive observational studies, case studies/series, evaluation studies, and accredited conference proceedings related to pandemic management will be included after assessing the quality. Also, reference checking of detected peer reviewed studies and hand searching of related journals will be conducted.

#### Exclusion criteria

Empirical studies, commentaries, letter to editor that did not mention health system resilience strategies for pandemic/epidemic management will be excluded.

#### Source of information and Search strategies

World Health Organization’s (WHO) six building blocks of health system,[3] were included as the key words. Boolean operators (“AND” or “OR”) and asterisk search operators will be applied. The complete search strategies for all databases are presented in a table. The WHO depository will be accessed for Health Systems policy analysis. To avoid duplication and for citation purposes, references will be collected from each database and stored in EndNote desktop version 20. The search will be restricted to literature related to the management of declared PHEIC after 2007 according to the IHR for global health security,[6]. Screening, reference checking of detected studies and hand searching of related journals were conducted from 30 ^th^ of May to 15 ^th^ of July 2022. Electronic databases (PUBMED, EMBASE, SCOPUS, Web of Science) and Google scholar search engine which were published after declaration of PHEIC will be used to describe the related management strategies and lesson learned. The relevant keywords under the main three themes listed in table 1 will be used to create a comprehensive search strategy. The search terms will be filtered by a combination of different key words, Medical Subject Headings (MeSH), Emtree terms, SCOPUS, and WOS search strategies as in the supplementary file 1.

**Table 1.** The applicable keywords and/or phrases under the main three themes.

|  |  |  |  |  |
| --- | --- | --- | --- | --- |
| Theme 1 |  | Theme 2 |  | Theme 3 |
| Health system |  | Resilience,[1-2] |  | Pandemic/PHEIC,[6] |
| Health system building blocks,[3]<br>“Service delivery” OR<br>“Health Resource*” OR<br>“Health workforce” OR<br>“Health information system*” OR “Health Product*” OR Vaccine*<br>OR Medicine* Or<br>Diagnostic* OR “Health Financing” OR “Health Leadership” OR “Health Governance” | AND | Resilien* OR Robust<br>OR Anti-fragil* OR<br>Adaptab* OR<br>Transformat* OR<br>Preserv* | AND | “Public Health Emergencies of International Concerns” OR<br>“PHEIC” OR<br>Pandemic OR<br>Outbreak OR<br>Epidemic OR<br>COVID-19 OR<br>Ebola OR H1N1 OR<br>“Influenza Pandemic” OR Zika<br>OR Poliomyelitis |

#### Condition and domain being studies

Global implementation of health system resilience strategic analysis combined with PHEIC management will be studied to provide recommendation for future pandemic preparedness.

#### Population

Global studies which describe health system resilience strategies will be taken as the study population.

#### Intervention(s)/ Exposure(s)

Global health system resilience enhancing strategies for the management of pandemics that were declared as PHEIC will be included to identify opportunities and challenges and provide recommendations towards successful management of public health emergencies.

#### Context

Global Health system resilience-enhancing strategies for the management of PHEIC and implementation will be assessed.

#### Main outcome(s)

Application of resilience-enhancing strategies for the improvement of managing PHEIC in all six areas of the health system,[3] (including Governance/leadership, Healthcare financing, Healthcare service delivery, Heath workforce, Health information system and access to medicine/vaccine) towards better management of pandemic.

#### Additional outcome(s)

Success, opportunities, and challenges towards the management of PHEIC will be identified in view of constructing recommendations for the management of future pandemic situations. The impact of strong health system resilience-enhancing strategies on reducing morbidity and mortality due to proper management of pandemic

## Data Management

### Data extraction

The initial search was performed by reviewers using pre-determined search terms and strategies from chosen databases. After the removal of the duplications, the titles, abstracts, and keywords were screened for relevance and eligibility criteria. The two reviewers will then extract the data from the studies selected for inclusion using a predesignated extraction form. We will use a Microsoft Excel spreadsheet format for data extraction. It will be done by two research team members (RMNUR and NP) independently to reduce the bias. We also conduct a double check-up and verification of the extracted information by the supervisory senior author (YA). The data extraction form consists of the details about the Title of the study, author, publication year, Study question/ Objectives, study setting/country, study design, analysis methods, exposure/main results (strategic area of six building blocks),[3], challenges/weaknesses, success, and study limitations [Refer Supplementary file 2]. Two reviewers (RMNUR and RK/AS/NP/GS) will be independently coded and tabulated on the findings. Adapting from the WHO Six Building blocks, we use the six key areas of the Health system, as a guiding theme for the reporting of findings.

### Risk of bias (quality) assessment

All retrieved studies were initially imported into the Rayyan software to assist in removing duplicates. After removing the duplicates, it was shared among collaborators for independent screening of articles by title and abstract based upon eligibility criteria. A three-stage screening process will be used to eliminate non-relevant articles at the stage of title, abstract, and full-text screening. The studies which would have the two or more reviewers agreed on will be subjected to a full-text review. The two reviewers will independently review the full text of all eligible studies that meet the inclusion criteria and will be retained for the final synthesis (RMNUR and NP). All eligible retrieved articles will undergo a quality assessment process during the synthesis of results and will be done by four independent reviewers (YA, NUR, RK, AS). The two independent reviewers (RMNUR and NP) will use the Joanna Briggs Institute’s (JBI) critical appraisal checklist for the qualitative research assessment. When there will be a disagreement between the two reviewers, the senior team member (YA) and collaborators (CA, MSDW, TKT) will be engaged and a discussion along with the two reviewers will be made to resolve the differences.

### Strategies for data synthesis

Double check-up and verification of the extracted information will be done by the senior reviewers (YA, CA, MSDW, TKT). Two team members (RMNUR and NP/GS) will be independently coded and tabulated the findings on the thematic areas (6 core areas of the WHO health system) of the review and will map out common codes, concepts, and categories. Adapting from the WHO framework, we use the six core areas of the health system, as a guiding theme for the reporting of findings. The key findings will be collated under those core areas of the health system including Governance/leadership, Healthcare financing, Healthcare service delivery, Heath workforce, Health information system and access to medicine/vaccine. The investigators initially discussed the analytical themes/framework independently and then collectively as a group to minimize the bias. Thus, the senior author (YA) and senior collaborators (CA, MSDW, TKT) cross-validate and resolve any discrepancies. The four main team members (RMNUR, RK, AS, NP) will also be regrouped as successes, challenges, and strategies to realize the management of the pandemic. All the reviewers will initially generate the analytical themes independently and then collectively as a group to minimize the bias. This framework offers a detailed list of likely factors that could contribute to the possible strategies for the attainment of six building blocks for enhancing the health system for pandemic management. The senior team member (YA) built upon the five team members’ work by categorizing and synthesizing applicable and emergent codes into final relevant themes, cross-validate all synthesized findings, and resolving any discrepancies.

### Analysis of subgroups or subsets

The successes and challenges of health system resilience strategies on the management of pandemics (PHIC) with regard to the context of achieving impacts, including improved health outcomes, equity, social and financial risk protection, responsiveness, and efficiency, which contribute to strengthening health systems to provide sustainable quality, efficient and effective health services will be assessed.

### Patient and Public involvement

None

## Discussion

Health system resilience is key to coping with catastrophic events, such as the economic crisis and COVID-19 pandemic. The mapping of available literature towards the application of resilience-enhancing strategies with existing frameworks needs to be assessed to identify the management gaps and give valuable recommendations from the lessons learnt from the global pandemic to improve the level of preparedness and response to similar public health emergencies in the future.

## Conclusions

A protocol for a global review of health system resilience for pandemic management is described. This review will add to the body of knowledge about health system enhancing research and policy formulation.

## Data Availability

All data produced in the present study are available upon reasonable request to the authors

https://www.crd.york.ac.uk/prospero/display_record.php?RecordID=352612

## Abbreviations

COVID19: Novel coronavirus disease-19;
IHR: International Health Regulations;
PHEIC: Public Health Emergencies of International Concerns;
WHO: World Health Organization.

## Language

English

## Country

Australia & Sri Lanka

## Stage of review

Review ongoing

## Declarations

## Ethics and dissemination

Databases that require an institutional license will be accessed through the library at Postgraduate Institute of Medicine (PGIM), University of Colombo Sri Lanka, and The University of Queensland Australia. The findings will be presented at conferences at The University of Queensland in Australia, PGIM, University of Colombo Sri Lanka, and other locations around the world. Finally, the review’s findings will be published in a peer-reviewed international journal and will be disseminated to global communities.

Ethics approval and consent to participate: Not applicable

## Acknowledgement

- PGIM, University of Colombo, Sri Lanka
- Ministry of Health, the Government of Sri Lanka
- School of Public health, Faculty of Medicine, The University of Queensland, Australia

## Funding

This research received no specific grant from any funding agency in the public, commercial or not-for-profit sectors.

## Author contribution

Conceptualization: RMNUR, YA & CA, Draft the protocol: RMNUR; Supervision: YA; Review the protocol and feedback: YA, CA, MSDW, RK, AE, TT, NP, GS, RR, MF; Editing: RMNUR

## Conflict of Interests

The authors declare that they have no competing interests.

## Notes

### Competing Interest Statement

The authors have declared no competing interest.

### Funding Statement

This study did not receive any funding

### Summary of Updates

Inclusion criteria We intend to include all studies in English with any peer reviewed articles under the World Health Organization (WHO) six building blocks of health system that adhered to resilience enhancing strategies for the management of PHEIC after establishment of IHR since managing PHEIC after the establishment of IHR in 2007

## References

1. Thomas S., Sagan A., Larkin J., Cylus J., Figueras J., & Karanikolos M. POLICY BRIEF 36 Strengthening healthsystems resilience: Key concepts and strategies. World Health Organization. 2020.

2. Barasa E. W., Cloete, K., Gilson L. From bouncing back, to nurturing emergence: reframing the concept of resilience in health systems strengthening, Health Policy and Planning, Volume 32, Issue suppl_3, November 2017, Pages iii91–iii94, https://doi.org/10.1093/heapol/czx118

3. WHO. Monitoring the building blocks of health system: a handbook of indicators and their measurement strategies. 2010. WHO Document Production Services, Geneva, Switzerland. https://apps.who.int/iris/bitstream/handle/10665/258734/9789241564052-eng.pdf

4. Barasa E, Mbau R, Gilson L. What Is Resilience and How Can It Be Nurtured? A Systematic Review of Empirical Literature on Organizational Resilience. Int J Health Policy Manag. 2018 Jun 1;7(6):491–503. doi: 10.15171/ijhpm.2018.06. PMID: 29935126; PMCID: PMC6015506.

5. Bozorgmehr, K., Saint, V., Kaasch, A., Stuckler, D., & Kentikelenis, A. COVID and the convergence of three crises in Europe. The Lancet Public Health, 5(5), e247–e248. 2020. https://doi.org/10.1016/s2468-2667(20)30078-5

6. Wilder-Smith, A., & Osman, S. Public health emergencies of international concern: a historic overview. Journal of Travel Medicine, 27(8), 2020. https://doi.org/10.1093/jtm/taaa227

7. Legido-Quigley, H., Mateos-García, J. T., Campos, V. R., Gea-Sánchez, M., Muntaner, C., & McKee, M. The resilience of the Spanish health system against the COVID-19 pandemic. The Lancet Public Health, 5(5), e251–e252. 2020. https://doi.org/10.1016/s2468-2667(20)30060-8

8. Ager, A., Annan J., Panter-Brick C. Resilience: From Conceptualization to Effective Intervention. 2013. http://www.cpcnetwork.org/resource/resilience-from-conceptualization-to-effective-intervention/

9. Formana R. A. R., McKee M., Mossialosad E. 12 Lessons learned from the management of the coronavirus pandemic. Health Policy, 124, 577–580. 2020. Elsevier

10. Castleden M., M. M., Murray V, Leonardi G. Resilience thinking in health protection. Journal of Public Health, Vol. 33, (No. 3), pp. 369–377. 2011. https://doi.org/0.1093/pubmed/fdr027|

11. Saulnier, D. D., Blanchet, K., Canila, C., Cobos Munoz, D., Dal Zennaro, L., de Savigny, D., Durski, K. N., Garcia, F., Grimm, P. Y., Kwamie, A., Maceira, D., Marten, R., Peytremann-Bridevaux, I., Poroes, C., Ridde, V., Seematter, L., Stern, B., Suarez, P., Teddy, G., … Tediosi, F. A health systems resilience research agenda: moving from concept to practice. BMJ Glob Health, 6(8). 2021. https://doi.org/10.1136/bmjgh-2021-006779

