## Supplementary file [Search strategies] for "Health System Resilience-enhancing Strategies for Managing Public Health Emergencies of International Concerns (PHEIC): Success and Challenges A Systematic Review Protocol" (from another version of this preprint: version 2, 2022-08-22)

| **Database** | **Search Strategy** | **Document Results** |
| --- | --- | --- |
| PubMed | "Health System" OR "Service delivery" OR "Resources/Health workforce" OR "Health information systems" OR "Access to essential medicines" OR "Financing" OR "Leadership/governance" AND "Resilience" OR "Robust" OR "Antifragile" OR "Adaptable" OR "Transfermative" OR "Suppleness" OR "Pliancy" OR "Suppleness" OR "Ability to bounce back’ OR "Perseverance" OR "Workability" OR "Ductility" OR "Malleability’ AND "Pandemic" OR "Global" OR "Worldwide" OR "Universal" OR "Widespread" OR "Planetary" OR "Prevalent" OR "Pervasive" OR "Rampant" OR "Outbreak" OR "Epidemic" OR "COVID-19"  Mesh Terms: "Health Services"[Mesh] OR "Delivery of Health Care"[Mesh] OR "Health Resources"[Mesh] OR "Health Workforce"[Mesh] OR "Health Facilities"[Mesh] OR "Health Communication"[Mesh] OR "Healthcare Financing"[Mesh] OR "Medical Informatics"[Mesh] (Pandemic* OR Epidemic* OR Outbreak* OR “Public Health Emergencies of International Concerns”) AND (Management OR “Infection control” OR “Outbreak Control”) OR "Pandemics"[Mesh] OR "Disease Outbreaks/prevention and control"[Mesh] |  |
| Embase | (resilient:ti OR resilience:ti) AND (framework:ab,ti OR model:ab,ti OR program:ab,ti OR indicator:ab,ti OR index:ab,ti) AND (system:ab,ti OR organization:ab,ti OR organizational:ab,ti OR theory:ab,ti OR strategies:ab,ti) |  |
| Scopus | (*resilience* OR *resilient*) AND (*system* OR *model* OR *framework* OR *theory* OR *organiz** OR *organis** OR *concept* OR *program* OR *indicator*) AND (“*health system”* OR *“health sector”*) |  |
| WOS | (Resilience OR resilient) AND (System OR model OR framework OR theory OR organiz* OR organis* OR concept OR Strategies) AND (Epidemic OR “Epidemic” OR “health system” OR “health sector”) |  |
