## Supplementary material for "Health System Resilience-enhancing Strategies for Managing Public Health Emergencies of International Concerns (PHEIC): Success and Challenges A Systematic Review Protocol": Data extraction tool (from another version of this preprint: version 2, 2022-08-22)

**Supplementary file 2: data extraction tool for the systematic review**

| Title | Author /Year | Study question/ Objectives | Study area (Period) | Study design/type of study | Measurement/Analytical approach | Source of data | Main results including strategic areas (Six building blocks) | challenges/weaknesses, opportunities/Limitations | Comment |
| --- | --- | --- | --- | --- | --- | --- | --- | --- | --- |
